# Factors associated with congenital syphilis among women with gestational syphilis in a public health network in Bogotá, Colombia, 2017–2024: A case-control study

**DOI:** 10.64898/2026.11.10.26367783

**Authors:** Ana María Gómez Puentes, Narda María Olarte Escobar, Ismael Alberto Valderrama Márquez, Jorge Eliecer Castellanos Corredor, Ruth Liliana López Cruz, Javier Escobar-Pérez, Alejandro Rico Mendoza, Alexandra Porras Ramírez

## Abstract

**Background:** Congenital syphilis remains a preventable cause of perinatal morbidity and mortality, and its incidence in Colombia remains well above the elimination target. We aimed to identify maternal and prenatal care factors associated with congenital syphilis among women with gestational syphilis attended in a public health network serving socially vulnerable populations in Bogotá, Colombia.

**Methods:** We conducted a retrospective case-control study using surveillance and clinical records from the Subred Integrada de Servicios de Salud Sur E.S.E. for pregnancies ending between 2017 and 2024. Cases were neonates with congenital syphilis according to national surveillance criteria; controls were neonates without congenital syphilis born to mothers with gestational syphilis in the same network. Factors associated with congenital syphilis were identified using multivariable logistic regression with backward elimination.

**Results:** We included 743 mother–infant pairs (285 cases and 458 controls). Maternal diagnosis after 28 weeks of gestation (adjusted odds ratio 17.75; 95% confidence interval 11.81–26.69), absence of prenatal care compared with eight or more visits (12.74; 5.03–32.25) and reinfection during pregnancy (3.02; 1.73–5.29) were independently associated with congenital syphilis, whereas a documented history of syphilis was associated with lower odds (0.45; 0.25–0.82). The model showed high discrimination (area under the receiver operating characteristic curve 0.860). Migrant status was associated with congenital syphilis in the crude analysis but not after adjustment.

**Conclusions:** Congenital syphilis in this setting was associated mainly with late maternal diagnosis and absent prenatal care, conditions that preclude timely benzathine penicillin treatment. Strengthening early prenatal screening, continuity of care and partner management may reduce vertical transmission in socially vulnerable urban populations.

## Introduction

Congenital syphilis (CS) remains one of the most preventable yet persistent infectious causes of perinatal morbidity and mortality worldwide. The World Health Organization (WHO) estimated 661,000 cases of CS in 2016, resulting in 355,000 adverse birth outcomes, including stillbirths, neonatal deaths, preterm births and low birth weight [1]. The burden of syphilis among women of reproductive age remains substantial [2], and the WHO validation criteria for elimination of mother-to-child transmission require a CS incidence of 50 or fewer cases per 100,000 live births, together with antenatal screening and treatment coverage of at least 95% [3].

In the Americas, the Pan American Health Organization reported 29,149 CS cases in 2020, 76% of them in Brazil [4]. In Colombia, the prevalence ratio of gestational syphilis doubled between 2019 and 2024, from 11.1 to 22.0 per 1,000 live births plus fetal deaths, and 1,097 CS cases were notified in 2024, an incidence of 2.5 per 1,000 live births plus fetal deaths, five times the elimination threshold [5]. Among these cases, 90.4% of maternal diagnoses occurred in the third trimester, 37.4% of mothers had no prenatal care and only half received at least one dose of treatment [5]. These figures indicate a systemic failure of early prenatal serological screening, a paradox given that benzathine penicillin prevents vertical transmission when administered appropriately and at least 30 days before delivery [6]. National clinical practice guidelines therefore recommend syphilis screening at the first prenatal visit, with repeat testing in each trimester and at delivery [7]. The biological basis for this window is well established: the ability of *Treponema pallidum* subsp. *pallidum* to cross the placental barrier increases from approximately 16 weeks of gestation onward, making early detection the critical intervention point [8].

CS is also re-emerging in high-income settings. The European Centre for Disease Prevention and Control reported 78 confirmed CS cases across the European Union and European Economic Area in 2023, with increases that paralleled rising syphilis notification rates among women [9]. In Eastern Europe, Bulgaria reported an incidence of 20 cases per 100,000 live births in 2016 and a 250% increase in case numbers between 2017 and 2019 [10]. David et al. framed CS in Switzerland as a marker of inequality, because cases concentrated where prenatal screening, treatment continuity and access to care failed [10]. These findings support the interpretation of CS as a sentinel event for inequities in access to timely prenatal and sexually transmitted infection care.

The Subred Integrada de Servicios de Salud Sur E.S.E. (Subred Sur) is a public health service network in southern Bogotá that integrates primary care units and referral hospitals serving the localities of Ciudad Bolívar, Usme, Tunjuelito and Sumapaz, among the lowest-income areas of the city, and attends approximately 6,000 deliveries annually [11]. Its catchment includes Venezuelan migrants and other socially vulnerable populations who may face compound barriers to prenatal care access and continuity [12]. Studies of CS risk factors have been conducted mainly in settings with different health-system arrangements and social contexts, including Brazil, China, Switzerland and the United States [13–18]. In Colombia, analytical evidence remains limited to a few settings, such as the Pacific coast [19] and a cohort evaluating a positive deviance strategy in Bogotá [20]. We aimed to identify maternal and prenatal care factors independently associated with CS over an 8-year period in this large, socially complex urban public health network.

## Materials and methods

### Study design and setting

We conducted a retrospective case-control study using routinely collected data from January 1, 2017, through December 31, 2024. The study is reported in accordance with the Strengthening the Reporting of Observational Studies in Epidemiology (STROBE) guidelines [21] (S1 Checklist). The Subred Sur is the sole public integrated health network in southern Bogotá, serving a population of approximately 1.3 million inhabitants; it provides obstetric care regardless of insurance status and is the primary referral network for migrants and other vulnerable populations. Data were obtained from the institutional communicable disease databases of the Early Warning team of the Subred Sur Health Risk Management Directorate, institutional electronic health records, cases notified to the Colombian national public health surveillance system (SIVIGILA) and the national birth and death registry (RUAF).

### Participants

All women with gestational syphilis confirmed according to the criteria of the Colombian National Institute of Health (INS) whose pregnancy ended at a Subred Sur facility during the study period were eligible. Pregnancies ending in out-of-facility delivery, spontaneous abortion, ectopic pregnancy or induced abortion were excluded.

### Case and control definitions

Cases were neonates diagnosed with CS according to INS national surveillance criteria [22], which are consistent with WHO definitions and complementary to the diagnostic standards of the US Centers for Disease Control and Prevention [23]: a case is confirmed when either an epidemiological criterion (neonate born to a mother with untreated or inadequately treated syphilis) or a laboratory criterion (neonatal nontreponemal antibody titer at least fourfold higher than the maternal titer at delivery) is met. Controls were neonates without CS born to mothers with confirmed gestational syphilis who delivered within the same network during the same period. All eligible controls identified over the eight-year study period were included, without sampling or matching, yielding an observed case-to-control ratio of 1:1.6. Because controls were not selected through matching, unconditional logistic regression was used for all analyses.

#### Relationship between exposures and the case definition

Under the INS surveillance definition, a neonate born to a mother with untreated or inadequately treated syphilis is classified as a case on epidemiological grounds alone. Maternal treatment adequacy (completion of benzathine penicillin at least 30 days before delivery) is therefore a component of the outcome definition rather than an independent exposure, and, by construction, all control mothers had received adequate treatment. For this reason, maternal treatment was not analysed as a risk factor; it is reported descriptively to characterise the care pathway, and no measure of association is presented for it. Gestational age at diagnosis is also related to the case definition, because diagnosis late in pregnancy shortens the interval available to complete treatment 30 days before delivery. It does not, however, determine case status: women diagnosed after 28 weeks may still complete treatment within this window before a term delivery, as occurred in 15.7% of controls. Late diagnosis was therefore retained as an exposure, and its estimate is interpreted in light of this structural relationship (see Limitations).

### Sample size

Sample size was determined by the total number of eligible records over the study period (census sampling). To assess whether this sample was sufficient, we estimated the minimum size required to detect an odds ratio of 2.0 with a two-sided α of 0.05, 80% power and a 1:2 case-to- control ratio, assuming an exposure prevalence of 16% among controls (Kelsey method) [24]; the required sample was 158 cases and 315 controls. Because the exposure prevalence was derived from the study controls, this calculation is post hoc and is reported only to show that the available sample exceeded the minimum requirement.

### Variables

The unit of analysis was the pregnant woman, because the exposures of interest (timeliness of diagnosis, treatment and access to prenatal care) are measured in the mother, although the outcome occurs in the neonate. Variables were organised into six domains: (1) sociodemographic: maternal age, migrant status (country of origin other than Colombia), educational level, health insurance status and locality of residence; (2) obstetric history: obstetric formula, prior documented syphilis, HIV infection and psychoactive substance use; (3) prenatal care: gestational week at first visit, total number of prenatal visits and psychosocial risk; (4) diagnosis: gestational age at maternal syphilis diagnosis (continuous and dichotomised at >28 vs. ≤28 weeks, the end of the gestational period in which vertical transmission most frequently occurs [22]) and reinfection during pregnancy, defined as a new serological rise compatible with active infection in a woman previously treated during the same pregnancy; (5) treatment: number of benzathine penicillin doses, completion at least 30 days before delivery and partner treatment; and (6) outcome: CS according to INS criteria. Prenatal care was categorised as none, 1–7 visits or eight or more visits, following the WHO recommendation of a minimum of eight contacts [25].

Adequate treatment was defined as completion of the full course of benzathine penicillin at least 30 days before delivery. Four variables were not entered into the multivariable model. Adequate benzathine penicillin treatment was not entered into the regression models because it is a component of the case definition; the complete separation observed (all 458 controls vs. 4 of 285 cases [1.4%]) is a direct consequence of this definitional overlap rather than an empirical finding. Partner treatment adequacy showed a bivariate association (crude OR 1.50, 95% CI 1.11–2.02; p = 0.009) but was not retained after backward elimination because it did not meet the retention criterion (p < 0.10) after adjustment for the remaining covariates; this likely reflects collinearity with reinfection, as both capture failure of the partner management pathway. Lack of health insurance was excluded as an upstream structural determinant acting through access to prenatal care, because its inclusion risked overadjustment for a mediator. Preterm delivery was excluded a priori on causal grounds, because *T. pallidum* infection can induce preterm labour, making temporal sequencing indeterminate in retrospective data.

### Statistical analysis

Categorical variables were described as frequencies and proportions and non-normally distributed continuous variables as medians with interquartile ranges (IQRs). Crude odds ratios (ORs) with 95% confidence intervals (CIs) were estimated by simple logistic regression, and group differences were assessed using Pearson’s chi-square test or the Mann–Whitney U test, as appropriate. Variables with a bivariate p < 0.20 and less than 10% missing data were candidates for multivariable analysis; this inclusive threshold was chosen to avoid the premature exclusion of variables that may act as confounders once other covariates are considered [26]. We used multivariable logistic regression with backward elimination (retention criterion p < 0.10), with final variable selection combining statistical thresholds and biological plausibility [26]. The confounding criterion was a change of 10% or more in any retained OR upon removal of a candidate variable. Multiplicative interaction terms between migrant status and all model covariates were evaluated; none were statistically significant (all p > 0.29). Model performance was assessed using the area under the receiver operating characteristic curve (AUC-ROC), sensitivity, specificity, McFadden’s pseudo-R², the Hosmer–Lemeshow goodness-of-fit test and the Brier score. All tests were two-sided with a significance level of 0.05. Analyses were conducted in Stata version 15.1 (StataCorp LLC, College Station, TX, USA) [27].

The multivariable model used complete-case analysis; 718 of 743 participants (96.6%) had complete data on all candidate variables and were included in the adjusted model. The 25 excluded participants (3.4%) had missing data primarily on educational level, which was excluded from candidate variable selection because it exceeded the 10% missing data threshold (missing in 12.3% of participants); the four variables retained in the final model each had less than 5% missing data. Given the small proportion excluded and the absence of systematic patterns suggesting informative missingness, complete-case analysis was considered appropriate.

### Ethics statement

This study was approved by the Comité Institucional de Ética en Investigación of the Subred Integrada de Servicios de Salud Sur E.S.E. (registry No. 314, Resolution 20255000-009679-1, April 16, 2025) and was classified as research without risk under Resolution 8430 of 1993 of the Colombian Ministry of Health [28]. It was conducted in accordance with the principles of the Declaration of Helsinki [29]. The ethics committee waived the requirement for individual informed consent because the study involved only secondary analysis of existing surveillance and clinical records. All records were anonymised with numerical codes before statistical analysis, in compliance with Colombian Law 1581 of 2012 on personal data protection [30], and no information allowing direct identification of mothers or neonates was used at any stage of the analysis. Data were accessed for research purposes from 01/05/2025 to 15/05/2026. The authors who extracted the data from clinical records had access to information that could identify individual participants during data collection; the dataset used for analysis contained no direct identifiers.

## Results

### Study population

Of 851 records screened, 793 met the criterion of delivery at a network facility (58 excluded for out-of-facility delivery), and 743 mother–infant pairs met full eligibility criteria (50 additional exclusions for spontaneous abortion, ectopic pregnancy or induced abortion), yielding 285 cases and 458 controls (Fig 1). Included deliveries occurred between January 2, 2017, and December 27, 2024.

**Fig 1.**
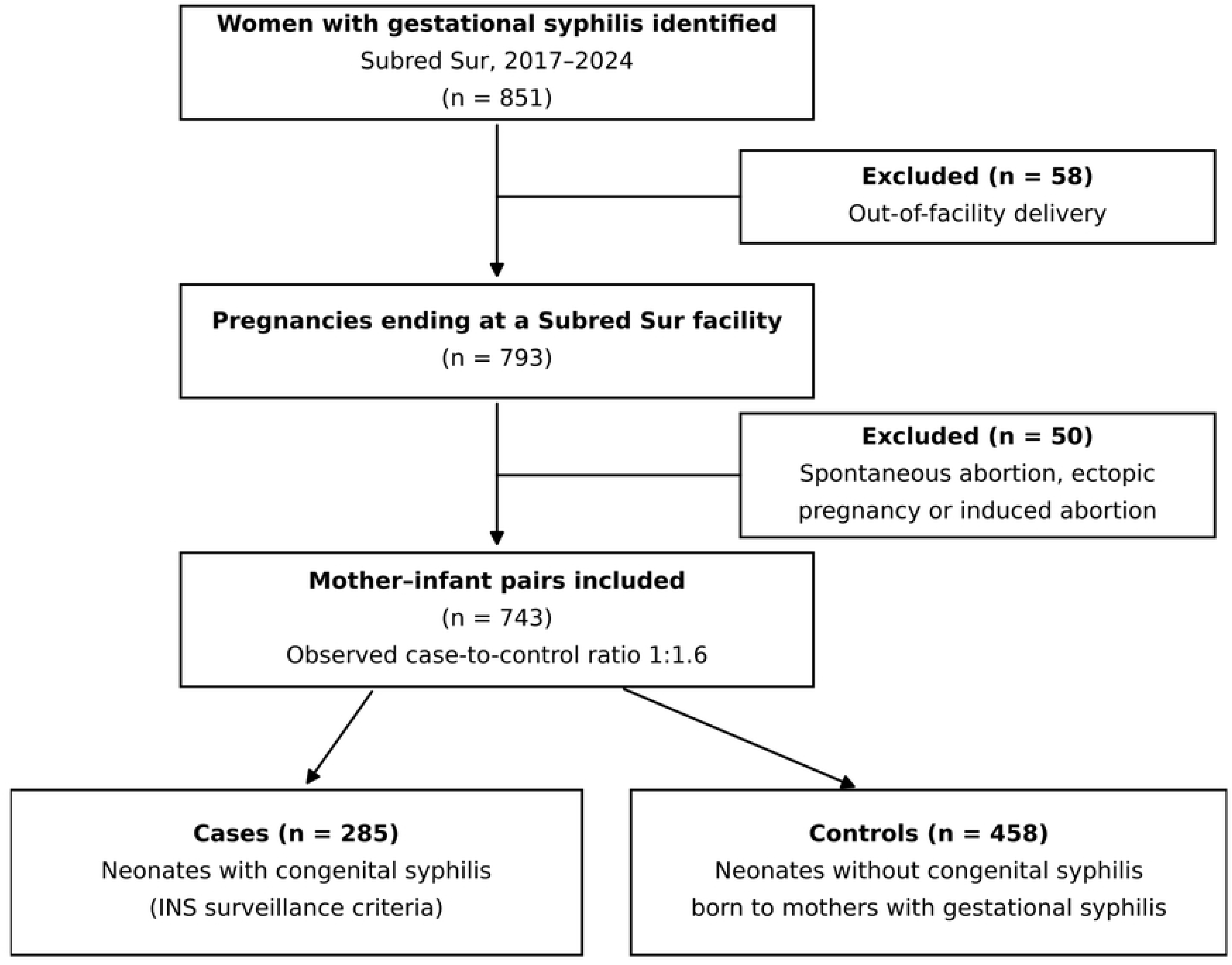
Flow diagram of participant selection. Women with gestational syphilis attended at the Subred Integrada de Servicios de Salud Sur E.S.E., Bogotá, Colombia, 2017–2024. INS, Colombian National Institute of Health.

Median maternal age was 23 years (IQR 20–27); cases were younger than controls (22 vs. 24 years; p = 0.027). Cases had higher proportions of international migrants (51.2% vs. 38.9%; p = 0.001), uninsured women (50.2% vs. 33.0%; p < 0.001), low educational attainment (primary education or less: 37.5% vs. 26.6%; p = 0.002) and high psychosocial risk (94.4% vs. 83.2%; p < 0.001) (Table 1). Prior syphilis history, substance use and reinfection are shown in Tables 1 and 2.

**Table 1.** Characteristics of women with gestational syphilis, by congenital syphilis status.

| **Characteristic** | **Cases (n = 285)** | **Controls (n = 458)** | **p** |
|---|---|---|---|
| **Sociodemographic** |  |  |  |
| Maternal age, median [IQR], years | 22 [20–27] | 24 [21–27] | 0.027 |
| International migrant | 146 (51.2) | 178 (38.9) | 0.001 |
| No health insurance | 143 (50.2) | 151 (33.0) | <0.001 |
| Low educational level (primary or none) | 107 (37.5) | 122 (26.6) | 0.002 |
| High psychosocial risk | 269 (94.4) | 381 (83.2) | <0.001 |
| **Medical history** |  |  |  |
| Prior syphilis history | 52 (18.2) | 113 (24.7) | 0.040 |
| Psychoactive substance use | 41 (14.4) | 81 (17.7) | 0.241 |
| **Prenatal care and diagnosis** |  |  |  |
| Gestational age at diagnosis, median [IQR], weeks | 35 [27–38] | 17 [10–25] | <0.001 |
| Late maternal diagnosis (>28 weeks) | 201 (70.5) | 72 (15.7) | <0.001 |
| Prenatal care visits<sup>a</sup> |  |  | <0.001 |
| None | 188 (66.0) | 168 (36.7) |  |
| 1–7 | 79 (27.7) | 208 (45.4) |  |
| ≥8 | 18 (6.3) | 80 (17.5) |  |
| Reinfection during pregnancy | 78 (27.4) | 157 (34.3) | 0.054 |
| **Treatment** |  |  |  |
| Adequate benzathine penicillin (completed ≥30 days before delivery)<sup>b</sup> | 4 (1.4) | 458 (100.0) | — |
| Inadequate partner treatment (<3 doses) | 157 (55.1) | 206 (45.0) | 0.009 |
| **Delivery outcomes** |  |  |  |
| Preterm delivery (<37 weeks) | 59 (20.7) | 26 (5.7) | <0.001 |
| Fetal death or stillbirth<sup>c</sup> | 28 (9.8) | 1 (0.2) | <0.001 |

Values are n (%) unless otherwise indicated. p values from Pearson's chi-square test for categorical variables and the Mann–Whitney U test for continuous variables. IQR, interquartile range.
<sup>a</sup> p value from the overall chi-square test across the three categories.
<sup>b</sup> Component of the case definition (Colombian National Institute of Health epidemiological criterion); by construction, all controls received adequate treatment. Reported descriptively; no test of association is presented.
<sup>c</sup> Ascertained by clinical record review.

**Table 2.** Crude odds ratios for factors associated with congenital syphilis.

| **Variable** | **Cases (n = 285)** | **Controls (n = 458)** | **Crude OR** | **95% CI** | **p** |
|---|---|---|---|---|---|
| **Prenatal care and diagnosis** |  |  |  |  |  |
| Late diagnosis (>28 vs. ≤28 weeks) | 201 (70.5) | 72 (15.7) | 12.83 | 8.97–18.35 | <0.001 |
| No prenatal care vs. ≥8 visits | 188 (66.0) | 168 (36.7) | 4.97 | 2.86–8.64 | <0.001 |
| 1–7 visits vs. ≥8 visits | 79 (27.7) | 208 (45.4) | 1.69 | 0.95–2.99 | 0.077 |
| Reinfection during pregnancy | 78 (27.4) | 157 (34.3) | 0.72 | 0.52–1.00 | 0.054 |
| **Medical history** |  |  |  |  |  |
| Prior syphilis history | 52 (18.2) | 113 (24.7) | 0.68 | 0.47–0.98 | 0.039 |
| **Sociodemographic** |  |  |  |  |  |
| International migrant | 146 (51.2) | 178 (38.9) | 1.65 | 1.23–2.23 | 0.001 |
| No health insurance | 143 (50.2) | 151 (33.0) | 2.05 | 1.51–2.77 | <0.001 |
| Low educational level (primary or none) | 107 (37.5) | 122 (26.6) | 1.66 | 1.21–2.27 | 0.002 |
| High psychosocial risk | 269 (94.4) | 381 (83.2) | 3.40 | 1.94–5.95 | <0.001 |
| **Treatment** |  |  |  |  |  |
| Inadequate partner treatment (<3 doses) | 157 (55.1) | 206 (45.0) | 1.50 | 1.11–2.02 | 0.009 |
| Adequate benzathine penicillin<sup>a</sup> | 4 (1.4) | 458 (100.0) | — | — | — |
| **Delivery outcomes** |  |  |  |  |  |
| Preterm delivery (<37 weeks)<sup>b</sup> | 59 (20.7) | 26 (5.7) | 4.34 | 2.66–7.07 | <0.001 |

Values are n (%). Crude odds ratios estimated by simple logistic regression. Variables with p < 0.20 and less than 10% missing data were candidates for multivariable analysis. CI, confidence interval; OR, odds ratio.
<sup>a</sup> Not estimated: component of the case definition (all controls received adequate treatment by construction).
<sup>b</sup> Not entered into the multivariable model because preterm delivery may be a consequence of fetal infection (see Methods).

### Prenatal care and timing of diagnosis

Late maternal syphilis diagnosis (>28 weeks of gestation) was present in 70.5% of cases vs. 15.7% of controls. Median gestational age at maternal diagnosis was 35 weeks for cases vs. 17 weeks for controls (p < 0.001). No prenatal care was reported by 66.0% of cases vs. 36.7% of controls, while adequate prenatal care (eight or more visits) was documented in only 6.3% of cases vs. 17.5% of controls (p < 0.001). Preterm delivery occurred in 20.7% of cases vs. 5.7% of controls (p < 0.001). Fetal death or stillbirth affected 9.8% of case pregnancies (n = 28) compared with 0.2% of controls (n = 1; p < 0.001). Complete bivariate results are presented in Tables 1 and 2.

### Benzathine penicillin treatment

Adequate benzathine penicillin treatment (completed at least 30 days before delivery) was documented in 4 of 285 case mothers (1.4%) and, as required by the case definition, in all 458 control mothers. This variable is presented descriptively and was not included in the regression models.

### Multivariable model

The final multivariable model included 718 complete cases and four covariates, all of which were independently associated with CS (Table 3). Late maternal diagnosis after 28 weeks of gestation showed the strongest association (aOR 17.75, 95% CI 11.81–26.69; p < 0.001).

**Table 3.** Multivariable logistic regression model of factors associated with congenital.

| **Variable** | **aOR** | **95% CI** | **p** |
|---|---|---|---|
| Maternal diagnosis >28 weeks of gestation (vs. ≤28 weeks) | 17.75 | 11.81–26.69 | <0.001 |
| Prenatal care (reference: ≥8 visits) |  |  |  |
| None | 12.74 | 5.03–32.25 | <0.001 |
| 1–7 visits | 1.74 | 0.87–3.49 | 0.120 |
| Reinfection during pregnancy (vs. none) | 3.02 | 1.73–5.29 | <0.001 |
| Documented prior syphilis (vs. none) | 0.45 | 0.25–0.82 | 0.010 |

n = 718 (complete-case analysis). Backward elimination (retention p < 0.10). AUC-ROC 0.860; sensitivity 73.9%; specificity 84.6%; McFadden pseudo-R² 0.323; Hosmer–Lemeshow χ² 15.03, df 6, p = 0.020; Brier score 0.145 (null model 0.236; Brier skill score 0.39). aOR, adjusted odds ratio; AUC-ROC, area under the receiver operating characteristic curve; CI, confidence interval.

Absence of prenatal care was associated with nearly 13-fold higher odds of CS relative to eight or more visits (aOR 12.74, 95% CI 5.03–32.25; p < 0.001), whereas 1–7 visits showed an intermediate, non-significant association (aOR 1.74, 95% CI 0.87–3.49; p = 0.120). Reinfection during pregnancy was associated with threefold higher odds (aOR 3.02, 95% CI 1.73–5.29; p < 0.001) despite a borderline inverse crude association (OR 0.72; p = 0.054), a pattern consistent with negative confounding by more intensive serological surveillance in reinfected women. Prior documented syphilis was independently associated with lower odds of CS (aOR 0.45, 95% CI 0.25–0.82; p = 0.010). Crude and adjusted ORs for the final-model variables are shown in Fig 2. Migrant status showed a significant crude association (OR 1.65, 95% CI 1.23–2.23; p = 0.001) that was attenuated after multivariable adjustment, consistent with mediation through access barriers.

**Fig 2.**
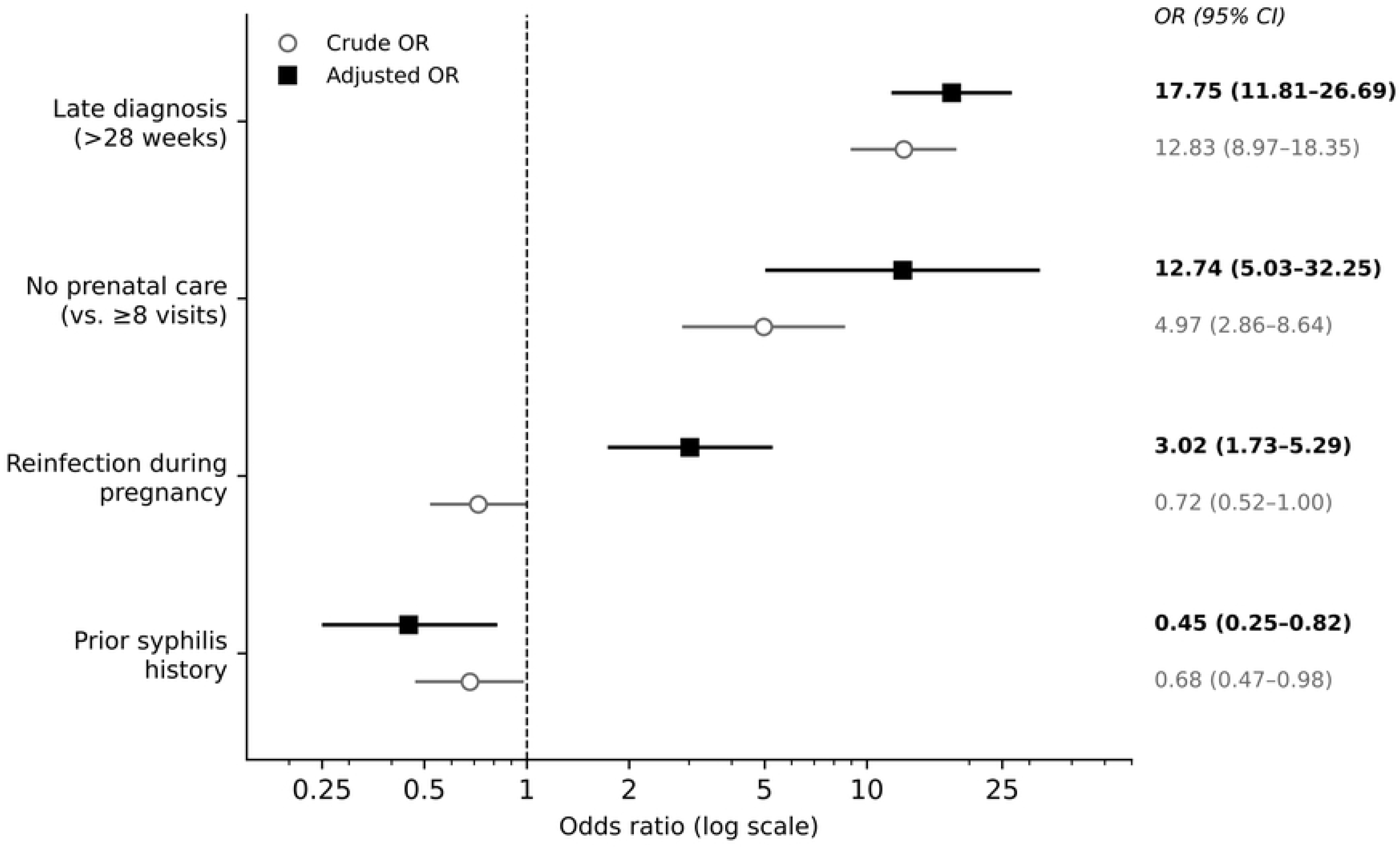
Crude and adjusted odds ratios for factors associated with congenital syphilis. Circles indicate crude odds ratios and squares adjusted odds ratios; horizontal lines indicate 95% confidence intervals on a logarithmic scale. The dashed vertical line indicates no association (OR = 1). PNC, prenatal care.

Model discrimination was high: AUC-ROC 0.860, sensitivity 73.9%, specificity 84.6% and McFadden pseudo-R² 0.323. The Hosmer–Lemeshow test yielded p = 0.020; because this test is sensitive to sample size and risk grouping, we interpreted it together with additional performance metrics [26]. The Brier score was 0.145 (null model Brier score 0.236; Brier skill score 0.39), indicating improved overall probabilistic performance relative to a null model.

## Discussion

The factors identified in this 8-year case-control study suggest that CS in a socially complex urban setting is associated primarily with failures in timely diagnosis and sustained access to prenatal care, rather than with limitations of the available therapeutic tools.

Late maternal diagnosis after 28 weeks of gestation showed the strongest association with CS (aOR 17.75). This finding is consistent with a cohort from Suzhou, China, in which diagnosis after 36 weeks was associated with markedly higher CS risk than first-trimester diagnosis [16], and with multicentre data from Switzerland, where diagnosis in the second or third trimester was strongly associated with a late first antenatal visit [17]. Although gestational thresholds differed across studies, the direction of association consistently shows that delayed diagnosis sharply reduces the opportunity for effective prevention before delivery. Part of the magnitude observed here, however, reflects the structural link between late diagnosis and the case definition described in the Methods. In Colombia, 90.4% of maternal diagnoses among CS cases occur in the third trimester [5], situating the Subred Sur findings within a broader national failure of early prenatal screening.

Absence of prenatal care was associated with nearly 13-fold higher odds of CS. In a systematic review of 14 studies, Pascoal et al. identified inadequate or late prenatal care and inadequate or late maternal treatment as significant risk factors for CS [14], and Etti et al. reported absent prenatal care, late diagnosis and incomplete treatment as the main determinants of vertical transmission in Fortaleza, Brazil [15]. In the Colombian Pacific coast, the absence of screening during prenatal care was also associated with CS [19]. The higher magnitude observed here likely reflects the extreme social vulnerability of the catchment population. Low prenatal care utilisation cannot be explained solely by supply-side barriers: structural and psychosocial factors, including fear of a sexually transmitted infection diagnosis, intimate partner coercion, substance use and institutional distrust, act as barriers that expanding service coverage alone cannot overcome [25].

Reinfection was associated with threefold higher adjusted odds of CS, despite appearing inversely associated in the bivariate analysis. This negative confounding is supported by S1 Table: reinfected women had lower rates of late diagnosis (24.7% vs. 43.9%), earlier gestational age at maternal diagnosis (median 18 vs. 26 weeks) and more prenatal visits (median 4 vs. 0; all p < 0.001), reflecting closer serological surveillance in this subgroup. After adjustment for these care-access factors, reinfection emerged as an independent marker of CS risk. The Nascer no Brasil study also identified failures in maternal and partner treatment as key components of vertical transmission [13]. These findings underscore the need for systematic, active partner management and contact tracing.

Prior syphilis history was associated with 55% lower odds of CS (aOR 0.45). This likely reflects differential clinical pathways: previously identified women enter subsequent pregnancies already flagged in the health record, enabling earlier serological screening and diagnosis at earlier gestational stages. Pascoal et al. likewise identified a prior history of adequately treated syphilis as a protective factor [14]. Systematic documentation of sexually transmitted infection history and differentiated monitoring protocols for women with prior syphilis are low-cost interventions that warrant evaluation [22].

Only 4 of 285 case mothers (1.4%) had documented adequate treatment before delivery. Because treatment adequacy is part of the surveillance case definition, this proportion describes the care pathway of cases rather than providing an independent estimate of treatment effect. Its relevance lies in what preceded it: the conditions that precluded timely treatment, namely late diagnosis and absent prenatal care, were the factors most strongly associated with CS in this population, which is consistent with the high efficacy of benzathine penicillin when administered at least 30 days before delivery [6,23]. Under the INS definition, the four adequately treated case mothers would have been classified through the laboratory criterion, and may represent treatment failure or reinfection close to delivery. Insurance status showed a crude association (OR 2.05) but was excluded from the model as an upstream determinant of access to prenatal care; in this institutional context, it also reflects differential ascertainment between public and contributory- regime networks.

Preterm delivery (20.7% of cases vs. 5.7% of controls) was not modelled as a risk factor, because *T. pallidum* infection can cause placental inflammation, fetal growth restriction and preterm labour, so that preterm birth may represent a consequence of uncontrolled fetal infection rather than an antecedent exposure. It is better interpreted as a clinical marker of the severity of untreated vertical transmission, as is the frequency of fetal death or stillbirth among cases (9.8% vs. 0.2% of controls). Preterm birth is one of the main adverse birth outcomes attributed to untreated maternal syphilis worldwide [1]. A comparison of the present findings with previous studies of CS risk factors is provided in S2 Table.

Migrant status was associated with CS in the crude analysis, but the association was attenuated after adjustment, a pattern also described in China [16]. This suggests that migration does not directly increase the risk of vertical transmission but acts through conditions associated with social vulnerability and access barriers, such as late diagnosis and absent prenatal care. Studies in Switzerland documented a high proportion of foreign-born mothers among CS cases [17], and in the United States public insurance has been associated with a higher frequency of syphilis during pregnancy, reflecting structural inequalities in prenatal access [31].

These findings collectively point to a single structural gap: the most vulnerable women access care late, intermittently or not at all. Fragmented care across institutions also means that prior syphilis history, previous serological titres and partner treatment records are often unavailable at the point of care. Universal first-trimester syphilis screening, with mandatory repeat testing in the third trimester for migrants, uninsured women and women without prior prenatal care, may be the most effective programmatic priority [6,7,23,25]. Systematic partner management could be protocolised as a condition for case closure [13,15], and interoperable obstetric records would enable differentiated follow-up of women with prior sexually transmitted infections, a low-cost intervention that warrants evaluation [22].

### Limitations

As a retrospective study, findings depend on the completeness of clinical records, which may be compromised when women received care at multiple institutions, potentially leading to underestimation of prenatal contacts and treatment episodes. Several exposures are structurally related to the outcome definition. The INS criterion prioritises surveillance sensitivity over diagnostic specificity and classifies neonates of untreated or inadequately treated mothers as cases, so some neonates classified on epidemiological linkage alone may represent probable rather than microbiologically confirmed CS. For the same reason, maternal treatment adequacy could not be evaluated as a risk factor, and associations for variables that determine the opportunity to treat, particularly late diagnosis, are likely to be larger than they would be under a microbiological case definition. This partial incorporation of exposure into the outcome may therefore inflate the magnitude, although not the direction, of the association observed for late diagnosis. The distribution of cases by classification criterion was not available, which precluded a sensitivity analysis restricted to laboratory-confirmed cases. In selected atypical cases, complementary molecular tools such as digital PCR and next-generation sequencing can support diagnostic confirmation, as recently shown in a CS case from the same institution [32].

Surveillance bias may affect reinfection detection, because women with more prenatal contacts have more opportunities for repeat serological testing, potentially leading to overestimation of reinfection prevalence in the better-monitored subgroup. The adjusted model used complete-case analysis and excluded 25 participants with missing data. Information on income, employment, intimate partner violence, social support and housing conditions was not available, and these factors may act as residual confounders. Finally, results are specific to this public network and may not represent CS epidemiology across the whole city; however, the structural risk patterns identified, including the mediating role of migration through access barriers and the concentration of vulnerability in public networks, may inform prevention strategies in comparable urban settings with high migration pressure.

## Conclusions

Late gestational syphilis diagnosis, absent prenatal care and reinfection were the factors most strongly associated with CS in this high-vulnerability urban public network over eight years, while a prior history of syphilis was independently associated with lower odds of CS. CS persistence reflects systemic failures in diagnostic timeliness and continuity of prenatal care rather than the absence of effective therapeutic tools. Strategies likely to reduce CS in socially vulnerable urban populations include early capture of pregnant women, universal first-trimester syphilis screening with third-trimester repetition, timely benzathine penicillin treatment and comprehensive management of sexual partners.

## Data Availability

The data underlying this study cannot be shared publicly because they contain sensitive clinical and epidemiological surveillance information on pregnant women and neonates, and their disclosure is restricted by Colombian law and by the conditions of approval of the institutional ethics committee. De-identified data are available upon reasonable request to the Subred Integrada de Servicios de Salud Sur E.S.E. (contact) for researchers who meet the criteria for access to confidential data.

## Acknowledgments

The authors thank the data management team of the Subred Integrada de Servicios de Salud Sur E.S.E. for their support, and Claudia Milena Guacaneme Parra and Zulay Astrid Bustos Murillo for assistance in extracting data from clinical records. Artificial intelligence tools (Claude, Anthropic, San Francisco, CA, USA) were used for manuscript language editing and literature search; all statistical analyses were conducted by the authors in Stata version 15.1. The authors take full responsibility for the content of the manuscript.

## Supporting information

**S1 Checklist. STROBE checklist for case-control studies.** Completed checklist indicating the page and line numbers where each item is reported.

**S1 Table. Prenatal care and diagnosis characteristics according to reinfection status during pregnancy.** Late diagnosis, gestational age at diagnosis and number of prenatal visits in women with and without reinfection (n = 743).

**S2 Table. Comparison of studies of risk factors for congenital syphilis.** Design, setting, period, sample and main associated factors reported in previous studies and in the present study.

## Notes

### Competing Interest Statement

The authors have declared no competing interest.

### Clinical Trial

N/A. This is a retrospective observational case-control study, not a clinical trial or prospective interventional study.

### Author Declarations

This study was approved by the Comité Institucional de Ética para la Investigación (Institutional Research Ethics Committee) of the Subred Integrada de Servicios de Salud Sur E.S.E., Bogotá, Colombia (protocol registry No. 314 approval letter 20255000-009679-1, April 16, 2025). The study was classified as research without risk under Resolution 8430 of 1993 of the Colombian Ministry of Health, as it consisted exclusively of a retrospective review of anonymized surveillance and clinical records individual informed consent was therefore not required (Article 16).

## References

1. Korenromp EL, Rowley J, Alonso M, Mello MB, Wijesooriya NS, Mahiané SG, et al. Global burden of maternal and congenital syphilis and associated adverse birth outcomes— Estimates for 2016 and progress since 2012. PLoS One. 2019;14(2):e0211720. doi: 10.1371/journal.pone.0211720

2. Newman L, Rowley J, Vander Hoorn S, Wijesooriya NS, Unemo M, Low N, et al. Global estimates of the prevalence and incidence of four curable sexually transmitted infections in 2012 based on systematic review and global reporting. PLoS One. 2015;10(12):e0143304. doi: 10.1371/journal.pone.0143304

3. World Health Organization. Global guidance on criteria and processes for validation: elimination of mother-to-child transmission of HIV, syphilis and hepatitis B virus. Geneva: World Health Organization; 2021.

4. Pan American Health Organization. Elimination of mother-to-child transmission of HIV and syphilis in the Americas [Internet]. Washington (DC): PAHO; 2021 [cited 2026 Oct 26]. Available from: https://www.paho.org/en/topics/elimination-mother-child-transmission-hiv-and-syphilis

5. Pérez Clavijo CL. Informe de evento 2024: sífilis gestacional y sífilis congénita [Internet]. Bogotá: Instituto Nacional de Salud; 2025 [cited 2026 Oct 26]. Available from: https://www.ins.gov.co/buscador-eventos/Informesdeevento/S%C3%8DFILIS%20CONG%C3%89NITA%20Y%20GESTACIONAL%20INFORME%20DE%20EVENTO%202024.pdf

6. World Health Organization. WHO guideline on syphilis screening and treatment for pregnant women. Geneva: World Health Organization; 2017.

7. Ministerio de Salud y Protección Social, Fondo de Población de las Naciones Unidas (UNFPA). Guía de práctica clínica basada en la evidencia para la atención integral de la sífilis gestacional y congénita. Guía No. GPC-2014-41. Bogotá: Ministerio de Salud y Protección Social; 2014.

8. Berman SM. Maternal syphilis: pathophysiology and treatment. Bull World Health Organ. 2004;82(6):433–8.

9. European Centre for Disease Prevention and Control. Congenital syphilis. In: ECDC. Annual epidemiological report for 2023 [Internet]. Stockholm: ECDC; 2025 [cited 2026 Oct 26]. Available from: https://www.ecdc.europa.eu/en/publications-data/congenital-syphilis-annual-epidemiological-report-2023

10. David A, Posfay-Barbe KM, Aguiar Nogueira C, Toutous Trellu L. Congenital syphilis in Switzerland: a marker of inequality? A mini-review. Front Public Health. 2023;11:1265725. doi: 10.3389/fpubh.2023.1265725

11. Subred Integrada de Servicios de Salud Sur E.S.E. Informe de gestión 2023 [Internet]. Bogotá: Subred Sur; 2024 [cited 2026 Oct 26]. Available from: https://www.subredsur.gov.co/wp-content/uploads/2024/02/INFORMEDEGESTION_2023.pdf

12. García García DM, Mayorga JM. Asentamiento y segregación residencial en la migración sur-sur: migrantes venezolanos en Bogotá-Colombia. Bitácora Urbano Territ. 2023;33(2):91–106. doi: 10.15446/bitacora.v33n2.106141

13. Domingues RMSM, Leal MC. Incidência de sífilis congênita e fatores associados à transmissão vertical da sífilis: dados do estudo Nascer no Brasil. Cad Saude Publica. 2016;32(6):e00082415. doi: 10.1590/0102-311X00082415

14. Pascoal LB, Carellos EVM, Tarabai BHM, Vieira CC, Rezende LG, Salgado BSF, et al. Maternal and perinatal risk factors associated with congenital syphilis. Trop Med Int Health. 2023;28(6):442–53. doi: 10.1111/tmi.13881

15. Etti M, Neto ASL, Monteiro HS, Araújo MAL, Sousa GS, Castro MC. Determinants of congenital syphilis in Fortaleza, Brazil: a retrospective case-control study. PLOS Glob Public Health. 2023;3(12):e0002626. doi: 10.1371/journal.pgph.0002626

16. Wang Y, Wu M, Gong X, Zhao L, Zhao J, Zhu C, et al. Risk factors for congenital syphilis transmitted from mother to infant—Suzhou, China, 2011–2014. MMWR Morb Mortal Wkly Rep. 2019;68(10):247–50. doi: 10.15585/mmwr.mm6810a4

17. Alberto C, Wagner N, Fougère Y, Meyer Sauteur PM, Scherler G, Aebi-Popp K, et al. Syphilis in pregnant women and congenital syphilis from 2012 to 2021 in Switzerland: a multicentre, retrospective study. Swiss Med Wkly. 2024;154:3678. doi: 10.57187/s.3678

18. Kachikis A, Schiff MA, Moore K, Chapple-McGruder T, Arluck J, Hitti J. Risk factors associated with congenital syphilis, Georgia, 2008–2015. Infect Dis Obstet Gynecol. 2023;2023:3958406. doi: 10.1155/2023/3958406

19. Fuertes-Bucheli JF, Buenaventura-Alegría DP, Rivas-Mina AM, Pacheco-López R. Congenital syphilis prevention challenges, Pacific coast of Colombia, 2018–2022. Emerg Infect Dis. 2024;30(5):890–9. doi: 10.3201/eid3005.231273

20. Olarte NM, Valderrama IA, Castellanos JE, Raba AM, Jiménez JM, Mojica JA. Efecto de la desviación positiva en el diagnóstico y tratamiento de la sífilis gestacional, y prevención de la sífilis congénita: estudio de cohortes. Rev Colomb Obstet Ginecol. 2024;75(4). doi: 10.18597/rcog.4198

21. von Elm E, Altman DG, Egger M, Pocock SJ, Gøtzsche PC, Vandenbroucke JP. The Strengthening the Reporting of Observational Studies in Epidemiology (STROBE) statement: guidelines for reporting observational studies. PLoS Med. 2007;4(10):e296. doi: 10.1371/journal.pmed.0040296

22. Instituto Nacional de Salud. Protocolo de vigilancia en salud pública: sífilis gestacional y congénita. Version 07 [Internet]. Bogotá: Instituto Nacional de Salud; 2024. doi: 10.33610/CWUE8188

23. Workowski KA, Bachmann LH, Chan PA, Johnston CM, Muzny CA, Park I, et al. Sexually transmitted infections treatment guidelines, 2021. MMWR Recomm Rep. 2021;70(4):1–187. doi: 10.15585/mmwr.rr7004a1

24. Kelsey JL, Whittemore AS, Evans AS, Thompson WD. Methods in observational epidemiology. 2nd ed. New York: Oxford University Press; 1996.

25. World Health Organization. WHO recommendations on antenatal care for a positive pregnancy experience. Geneva: World Health Organization; 2016.

26. Hosmer DW Jr, Lemeshow S, Sturdivant RX. Applied logistic regression. 3rd ed. Hoboken (NJ): Wiley; 2013. doi: 10.1002/9781118548387

27. StataCorp. Stata statistical software: release 15. College Station (TX): StataCorp LLC; 2017.

28. Ministerio de Salud de Colombia. Resolución 8430 de 1993, por la cual se establecen las normas científicas, técnicas y administrativas para la investigación en salud. Bogotá: Ministerio de Salud; 1993.

29. World Medical Association. World Medical Association Declaration of Helsinki: ethical principles for medical research involving human participants. JAMA. 2025;333(1):71–4. doi: 10.1001/jama.2024.21972

30. Congreso de la República de Colombia. Ley Estatutaria 1581 de 2012, por la cual se dictan disposiciones generales para la protección de datos personales. Diario Oficial No. 48.587; 2012.

31. Gulersen M, Lenchner E, Eliner Y, Grunebaum A, Johnson L, Chervenak FA, et al. Risk factors and adverse outcomes associated with syphilis infection during pregnancy. Am J Obstet Gynecol MFM. 2023;5(6):100957. doi: 10.1016/j.ajogmf.2023.100957

32. Bossa-Castro AM, Olarte Escobar NM, Valderrama Márquez IA, Abril D, Fuentes L, Corredor-Rozo ZL, et al. Detection of congenital syphilis via digital PCR and next- generation sequencing, Colombia. Emerg Infect Dis. 2026;32(8):1341–4. doi: 10.3201/eid3208.251737

